# Deep-learning-based white matter lesion volume in CT is associated with outcome after acute ischemic stroke

**DOI:** 10.1101/2023.03.20.23287467

**Authors:** Henk van Voorst, Johanna Pitkänen, Laura van Poppel, Lucas de Vries, Mahsa Mojtahedi, Laura Martou, Bart J. Emmer, Yvo B.W.E.M. Roos, Robert van Oostenbrugge, Alida A Postma, Henk A. Marquering, Charles B.L.M. Majoie, Sami Curtze, Susanna Melkas, Paul Bentley, Matthan W.A. Caan, the MR CLEAN No-IV and CONTRAST consortium collaborators

## Abstract

**Background:** It remains unclear if deep-learning-based white matter lesion (DL-WML) volume can predict outcome after ischemic stroke.

**Purpose:** We aimed to develop, validate, and evaluate DL-WML volume in NCCT as a risk factor and IVT effect modifier compared to the Fazekas scale (WML-Faz) in patients also receiving EVT in an EVT-capable center.

**Methods:** A deep-learning model for WML segmentation in NCCT was developed and validated internally and externally. The volumetric correspondence of DL-WML volume per mL was reported relative to expert annotation with the intraclass correlation coefficient (ICC) and a Bland-Altman analysis reporting bias and limits of agreement (LoA). In a post-hoc analysis of the MR CLEAN No-IV trial, univariable and multivariable regression models were used to report (un)adjusted common odds ratios ([a]cOR) to associate DL-WML volume and WML-Faz with the occurrence of symptomatic-intracerebral hemorrhage (sICH) and 90-day functional outcome with the modified Rankin Scale (mRS).

**Results:** DL-WML volumes were comparable with those of the ground truth for both the internal test set (10/20(50%) male, age median:72[IQR:67-85], ICC mean:0.91 95%CI:[0.87;0.94];bias:-3mL LoA:[-12mL;7mL]) and the external test set (36/101(36%) male, age median:59[IQR:42-73], ICC mean:0.87 95%CI:[0.71;0.95];bias:-2mL LoA:[-11mL;7mL]). 516 patients from the MR CLEAN No-IV trial (291/516(56%) male, age median:71 IQR:[62-79],) were analyzed. Both DL-WML volume and WML-Faz were associated with sICH (DL-WML volume acOR:1.31 95%CI[1.08;1.60], WML-Faz acOR:1.53 95%CI[1.02;2.31]) and mRS (DL-WML volume acOR:0.84 95%CI[0.76;0.94], WML-Faz acOR:0.73 95%CI[0.60;0.88]). Only for the unadjusted analysis, WML-Faz was an IVT effect modifier (p=0.046), DL-WML was not (p=0.274).

**Conclusion:** DL-WML volume and WML-Faz had a similar relationship with functional outcome and sICH.

**Summary statement:** Deep-learning-based white matter lesion volume in non-contrast CT can substitute human ratings to prognosticate symptomatic intracerebral hemorrhage and functional outcome at 90-days in acute ischemic stroke patients.

**Key points:**

1. This was the first study to show that white matter lesion volume in non-contrast CT based on deep-learning segmentations (DL-WML volume) was associated with symptomatic intracerebral hemorrhages and a worse functional outcome.
2. Compared to the Fazekas scale, regression models using DL-WML volume had a similar fit to the data.
3. White matter lesion load might be associated with more symptomatic intracerebral hemorrhages if IVT was given before EVT.

## Introduction

The benefit of intravenous thrombolysis (IVT) before endovascular treatment (EVT) for large vessel occlusion acute ischemic stroke (AIS) in EVT-capable stroke centers remains unclear (1–6). An intracerebral hemorrhage (ICH) is a common complication after AIS that can be induced by IVT (7–10) and frequently results in severe neurologic disability and death (11,12). In patients with a high white matter lesion (WML) burden, ICH occurs more often and is aggravated by IVT (11,12). Therefore, accurate quantification of the WML burden might be used to guide safe administration of IVT in patients also eligible for EVT in an EVT-capable stroke center.

Currently, visual rating scores such as the Fazekas scale are used to grade WML (WML-Faz) in non-contrast CT (NCCT) or MRI (13) but suffer from imprecision and inter-rater variability(14). In a post-hoc analysis of the Chinese DIRECT-MT trial, randomizing patients for IVT before EVT in EVT-capable stroke centers, WML-Faz was associated with functional outcome while no association was found with ICH and effect modification of IVT for ICH or functional outcome (15). Thus, in patients with high WML-Faz scores, no increased rate of IVT-induced ICH was observed. Besides visual rating scales, several automated methods have been developed for WML segmentation in MRI and NCCT allowing for a fast and quantitative evaluation of WMLs (16,17). These automated WML segmentation methods for NCCT have only been analyzed for diagnostic accuracy relative to visual rating scales and based on spatial and volumetric correspondence compared to an expert-based ground truth segmentation (16,17). It remains unclear how such an automated segmentation method relates to patient outcomes and if it is a feasible alternative to a visual rating scale for WML. Finally, recent improvements in deep-learning led to superior performance over currently used automated methods but have not been considered for WML segmentation in NCCT (19).

We aimed to develop a state-of-the-art deep-learning model to segment WML in NCCT. We hypothesized that automatically extracted deep-learning-based WML (DL-WML) volume has a similar predictive association as WML-Faz with symptomatic or asymptomatic intracerebral hemorrhage (sICH/aICH) and functional outcome. Furthermore, we hypothesized that both DL-WML volume and WML-Faz result in more ICH and a worse functional outcome if IVT is given.

## Methods

### Data and study design

Three retrospective imaging databases were used. Data from two centers (A and B) were used to train, internally, and externally validate segmentations obtained through a nnUnet deep-learning model (19). For the center A-1 cohort, we included 118 patients from a previous multicenter study (17) and added 147 patients retrospectively collected (A-2 cohort). For the center B cohort, 101 patients with FLAIR and NCCT imaging acquired within 3 months were included (16). Subsequently, data from the MR CLEAN No-IV trial was used to compare the value of WMLs according to the Fazekas scale and automatically extracted DL-WML volume (6). The MR CLEAN No-IV was a randomized controlled trial for intravenous alteplase (0.9mg/kg) before EVT or direct EVT in EVT-capable stroke centers including patients between January 2018 and October 2020 (6). Patients from the MR CLEAN No-IV were included if a baseline NCCT was available **(n=516). Figure 1** and **Online supplement A** describe the data specifics and reuse of previously reported data.

**Figure 1:**
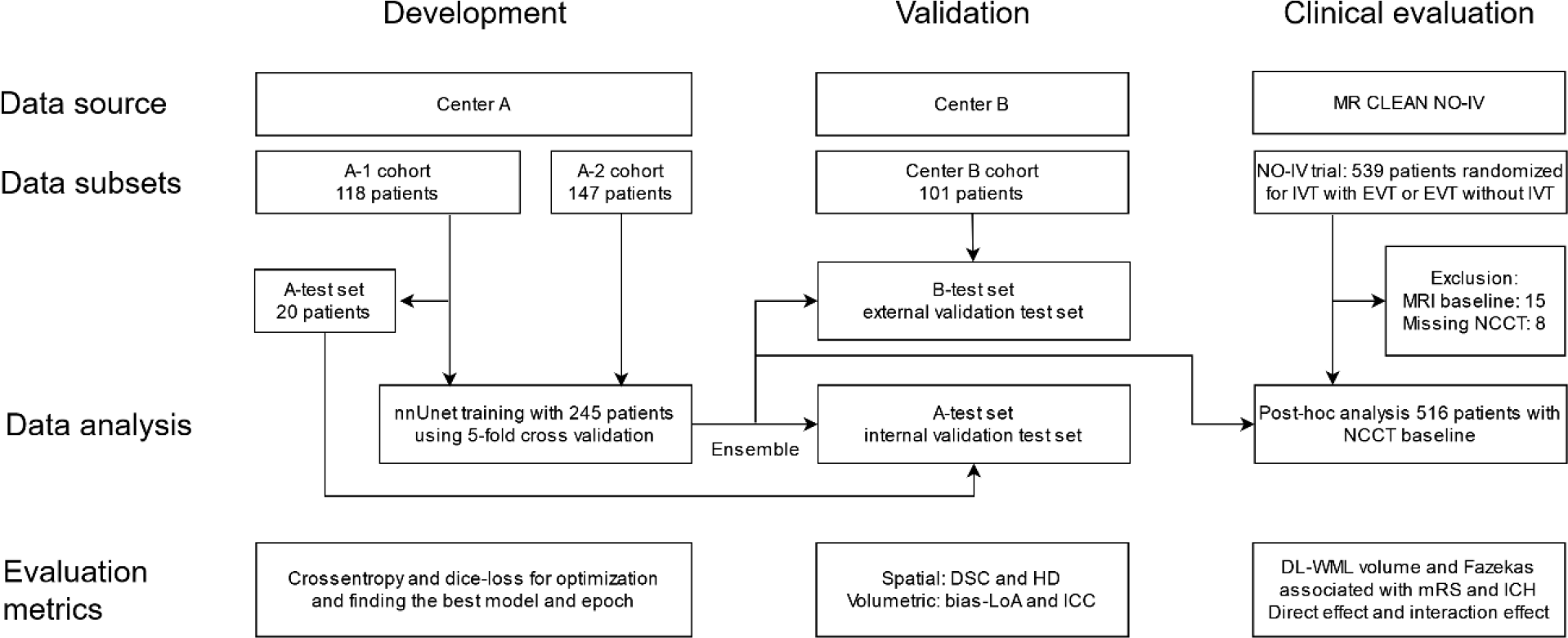
Visual methodology summary. nnUnet: no-new Unet, a deep-learning framework for segmentation. Missing NCCTs in the MR CLEAN No-IV were either not available in the database (n=3) or could not be converted to the correct format for automated analysis (n=5). DSC: Dice similarity coefficient. HD: Hausdorff distance. LoA: Limits of agreement. ICC: intra-class correlation coefficient. IVT: intravenous thrombolysis. NCCT: non-contrast CT. EVT: endovascular treatment. DL-WML volume: deep-learning-based white matter lesion volume segmented using the nnUnet framework. mRS: modified Rankin scale. ICH: intracerebral hemorrhage.

### Lesion annotation and data processing

In the center A cohort, ground-truth WMLs were segmented in NCCT by three experts (17). A single ground truth segmentation was created using the stapler algorithm which is based on a weighted majority voting between raters (20). WML-Faz was rated by any member of the independent core-lab. From the center A cohort (A1-train), we used 98 patients for training the nnUnet and 20 (center A-test set) for the internal validation test set. From the center A-2 cohort, 147 patients with single rater WML annotated slices were included for training (A2-train). For the center B data, NCCTs were co-registered to the FLAIR images using Elastix (21). Subsequently, WMLs in FLAIR were automatically segmented using medseg.ai (22) and manually adjusted by any of four raters (HV, LV, LP, and MM) using 3D-Slicer software, version 4.11 (23). All 101 patients from center B were used as an external validation test set (B-test).

### Lesion segmentation evaluation

The best nnUnet model was determined using the Dice similarity coefficient (DSC) for each of the 5 cross-validation folds. Evaluation metrics were reported based on a majority voting ensemble of these 5 models. To exclude false positive segmentations, we only considered WML segmentations inside a 10mm dilated mask of the ventricles (further described in **Online Supplement B)**. DL-WML segmentations were evaluated with the DSC and Hausdorff Distance (HD) in mm for spatial agreement, and volumetric correspondence using the bias and limits of agreement (LoA) in a Bland-Altman-plot and the two-way mixed effects intra-class correlation coefficient (ICC). The center A and B datasets were described with the total WML volume, average volume per WML, and the total WML count. Additionally, the NCCT quality was described with the signal-to-noise ratio (average voxel values in NCCT/standard deviation of background voxel values) and contrast-to-noise ratio ([average voxel values in WML – average background voxel values]/standard deviation of background voxel values) considering the entire brain tissue and a 5mm radius surrounding the WML as background.

### Post-hoc analysis MR CLEAN NO-IV

The primary outcome measure was functional outcome measured with the ordinal modified Rankin Scale (mRS) 90 days after AIS. Secondary outcome measures were functional independence (mRS≤2), mortality, sICH, and aICH according to the Heidelberg criteria (24). WML-Faz as a continuous variable and DL-WML volume were studied as independent variables. These WML measures were considered as risk factors for the outcome measures using binary and ordinal logistic regression models. Subsequently, a multiplicative interaction term of the WML measures with an intention-to-treat for IVT was introduced to the regression models to analyze treatment effect modification. Additional analyses for treatment effect modification were added in a per-protocol setting, excluding all patients that did not receive IVT and EVT according to the randomization. Model fit with DL-WML volume or the Fazekas scale was described with the likelihood ratio (LL). Odds ratios (ORs) with 95% confidence interval (95%CI) were reported for unadjusted univariate models and statistically adjusted multivariable models considering the following potential confounders: age, sex, pre-stroke mRS, collateral score, national institute of health stroke scale (NIHSS), Alberta stroke program early CT score (ASPECTS), IVT before EVT, time from onset of neurological deficit to arterial puncture, occlusion location, secondary hemostasis (INR: international normalized ratio), history of hypertension, history of diabetes mellitus, and previous cardiovascular disease consisting of peripheral artery disease, myocardial infarction, or ischemic stroke.

Baseline characteristics and WML measures were compared between the two treatment arms of the MR CLEAN No-IV with ANOVA, Kruskal-Wallis, and Chi-squared test for normal and non-normal distributed continuous, and categorical distributed baseline variables. **Online supplement A** describes the scoring of the imaging measures. All statistical analyses were performed in R version 3.6.3. For logistic regression analyses, missing values were imputed using multiple imputations for five datasets with the R-package MICE considering all potential confounders, outcome measures, and independent variables.

## Results

### Descriptive statistics

**Table 1** describes the characteristics of the NCCTs and the WML in the scans for the center A1-train set (55/98[56.1%] male sex, age median:76[IQR:66;85]), the A2-train set (66/147[44.8%] male sex, age median:78[IQR:68;85]), the center A-test set (10/20[50.0%] male sex, age median:72[IQR:67;85]), the center B-test set (36/101[35.6%] male sex, age median:59[IQR:42;73], and the included patients from the MR CLEAN No-IV trial (291/516[56.4%] male sex, age median:71[IQR:62;79]). All characteristics differed significantly across the datasets. The total WML volume and average volume per WML were higher in the center A datasets than in the B-test set. Furthermore, the number of lesions per patient was higher in the B-test set. Since the center A2-train set contained annotated slices and not volumes, the volume measures and number of WML are not directly comparable with other datasets. The signal-to-noise ratio was lower for the B-test set compared to other datasets. The contrast-to-noise ratio of the center A datasets was more negative than that of the B-test set. This indicates that in NCCTs from the B-test set WMLs were often in an area with similar Hounsfield Unit values inside the lesion as in the local background surrounding the WML. Patients in the B-test set and MR CLEAN No-IV had more often a thin slice (≤0.5mm) NCCT. **Table 2** describes the baseline characteristics of included and excluded patients from the MR CLEAN No-IV trial, per randomization arm and Fazekas scale sub-score. From the IVT before IVT arm 257 patients were included, 11 did not receive EVT or IVT. From the EVT without IVT arm 259 patients were included, in this arm, 28 patients did not receive EVT or did receive IVT. Patients with more severe WML according to the Fazekas scale were older, had a worse mRS before AIS, more often had a history of ischemic stroke, peripheral artery disease, diabetes mellitus, and hypertension, had a longer time from onset of neurological deficit to groin puncture, and had a larger DL-WML volume. **Online supplementary Table S1** describes the baseline characteristics between the included and excluded patients from the MR CLEAN No-IV trial.

**Table 1:** Dataset characteristics. WML: white matter lesions. NA: The MR CLEAN NO-IV dataset was used for clinical evaluation only; we did not have ground truth WML annotations for this dataset. IC: data from Imperial College. HUH: data from Helsinki University Hospital.

| **Dataset characteristics** | **A-1 training set** | **A-2 training set** | **A test set** | **B test set** | **MR CLEAN No-IV** |
| --- | --- | --- | --- | --- | --- |
| **Number of patients with NCCT** | 98 | 147 | 20 | 101 | 516 |
| **Age median (IQR)** | 76 (66;85) | 78 (68;85) | 72 (67;85) | 59 (42;73) | 71 (62;79) |
| **Male sex number (%)** | 55 (56.1%) | 66 (44.8%) | 10 (50%) | 36 (35.6%) | 291 (56.4%) |
| **Description of WML annotation used as ground truth** | NCCT volumes<br>3 raters | NCCT slices 1<br>rater | NCCT volumes<br>3 raters | NCCT volumes<br>once by any<br>of 3 raters | No annotations |
| **Total WML volume (mL)** | 11.4<br>(1.3;36.1) | 2.1<br>(1.1;4.6) | 17.1<br>(2.5;32.6) | 2.6 (0.6;9.4) | NA |
| **Number of WML** | 11(6;15) | 16 (6;32) | 13 (8;19) | 38 (12;71) | NA |
| **Average volume per WML** | 0.8<br>(0.2;2.2) | 0.1<br>(0.0;0.5) | 0.7<br>(0.3;3.0) | 0.1 (0.1;0.2) | NA |
| **Signal-to-noise ratio**<br>**(background=all brain tissue)** | 0.57<br>(0.53;0.59) | 0.62<br>(0.6;0.64) | 0.55<br>(0.51;0.60) | 0.38<br>(0.36;0.41) | 0.63<br>(0.59;0.68) |
| **Contrast-to-noise ratio**<br>**(background=5mm radius around ground truth)** | -0.57 (-0.79;-0.45) | -0.51 (-0.59;-0.40) | -1.13 (-1.26;-0.98) | -0.67 (-0.98;-0.47) | NA |
| **Slice thickness number (%)** |  |  |  |  |  |
| **≤0.5mm** | 0 (0.0%) | 0 (0.0%) | 0 (0.0%) | 19 (18.8%) | 276 (53.5%) |
| **0.5-1mm** | 0 (0.0%) | 146 (99.3%) | 0 (0.0%) | 59 (58.4%) | 70 (13.6%) |
| **1-5mm** | 7 (7.1%) | 1 (0.7%) | 1 (5.0%) | 13 (12.9%) | 169 (32.8%) |
| **>5mm** | 91 (92.9%) | 0 (0.0%) | 19 (95.0%) | 10 (9.9%) | 1 (0.2%) |

**Table 2:**
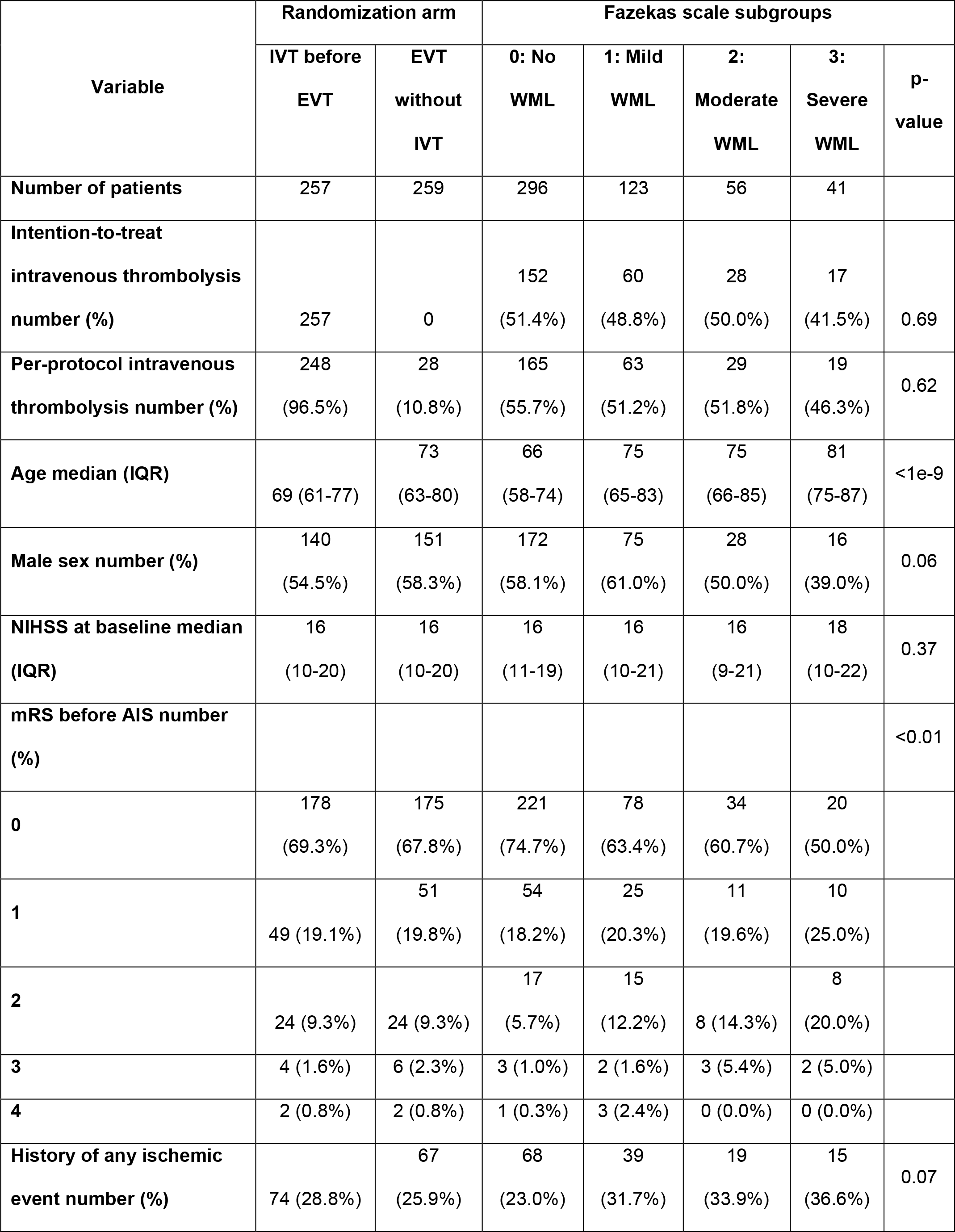
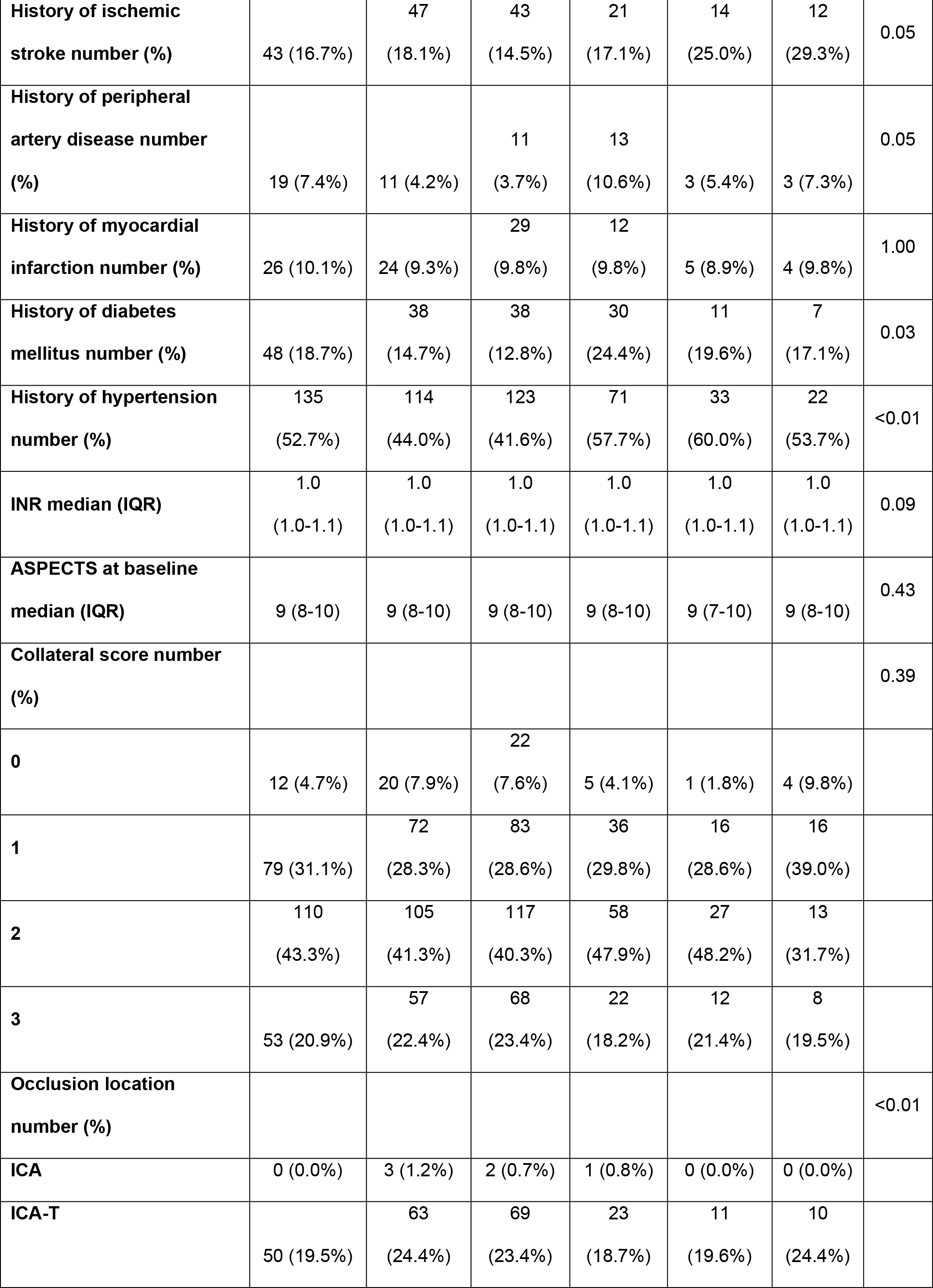
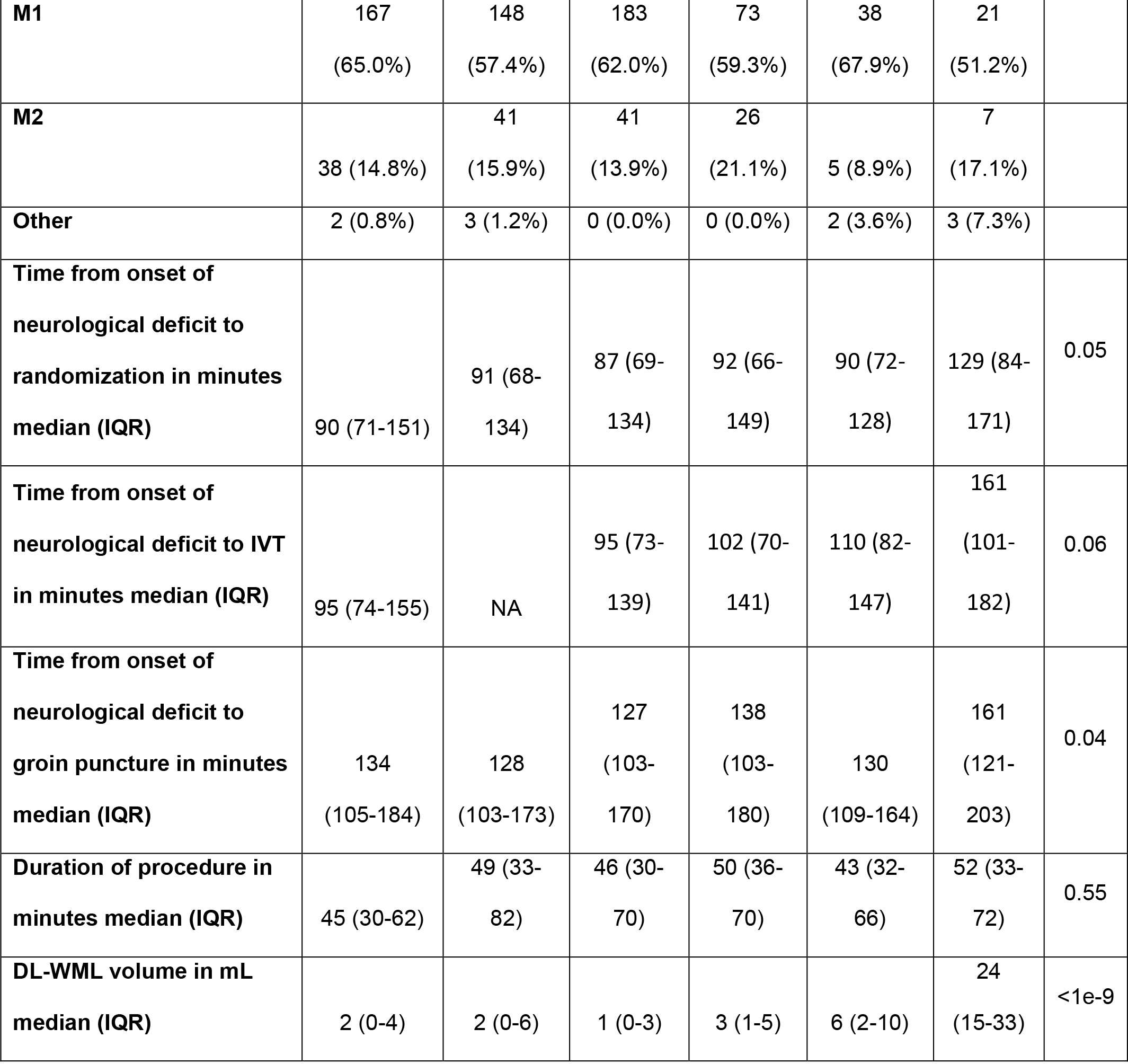
Baseline characteristics MR No-IV data used for this study. AIS: acute ischemic stroke. ASPECTS = Alberta Stroke Programme Early CT Score. IQR = interquartile range. mRS = modified Rankin Scale. NIHSS = National institute of health stroke. SD = standard deviation. INR: international normalized ratio (secondary hemostasis). DL-WML: deep-learning-based white matter lesion volume. ICA: internal carotid artery. ICA-T: ICA terminus. M1: M1 segment of the middle cerebral artery. M2: M2 segment of the middle cerebral artery. IVT: intravenous thrombolysis. NA: not available.

| Variable | Randomization arm |  | Fazekas scale subgroups |  |  |  |  |
| --- | --- | --- | --- | --- | --- | --- | --- |
|  | IVT before<br>EVT | EVT<br>without<br>IVT | 0: No<br>WML | 1: Mild<br>WML | 2:<br>Moderate<br>WML | 3:<br>Severe<br>WML | p-value |
| Number of patients | 257 | 259 | 296 | 123 | 56 | 41 |  |
| Intention-to-treat<br>intravenous thrombolysis<br>number (%) | 257 | 0 | 152<br>(51.4%) | 60<br>(48.8%) | 28<br>(50.0%) | 17<br>(41.5%) | 0.69 |
| Per-protocol intravenous<br>thrombolysis number (%) | 248<br>(96.5%) | 28<br>(10.8%) | 165<br>(55.7%) | 63<br>(51.2%) | 29<br>(51.8%) | 19<br>(46.3%) | 0.62 |
| Age median (IQR) | 69 (61-77) | 73<br>(63-80) | 66<br>(58-74) | 75<br>(65-83) | 75<br>(66-85) | 81<br>(75-87) | <1e-9 |
| Male sex number (%) | 140<br>(54.5%) | 151<br>(58.3%) | 172<br>(58.1%) | 75<br>(61.0%) | 28<br>(50.0%) | 16<br>(39.0%) | 0.06 |
| NIHSS at baseline median<br>(IQR) | 16<br>(10-20) | 16<br>(10-20) | 16<br>(11-19) | 16<br>(10-21) | 16<br>(9-21) | 18<br>(10-22) | 0.37 |
| mRS before AIS number<br>(%) |  |  |  |  |  |  | <0.01 |
| 0 | 178<br>(69.3%) | 175<br>(67.8%) | 221<br>(74.7%) | 78<br>(63.4%) | 34<br>(60.7%) | 20<br>(50.0%) |  |
| 1 | 49 (19.1%) | 51<br>(19.8%) | 54<br>(18.2%) | 25<br>(20.3%) | 11<br>(19.6%) | 10<br>(25.0%) |  |
| 2 | 24 (9.3%) | 24 (9.3%) | 17<br>(5.7%) | 15<br>(12.2%) | 8 (14.3%) | 8 (20.0%) |  |
| 3 | 4 (1.6%) | 6 (2.3%) | 3 (1.0%) | 2 (1.6%) | 3 (5.4%) | 2 (5.0%) |  |
| 4 | 2 (0.8%) | 2 (0.8%) | 1 (0.3%) | 3 (2.4%) | 0 (0.0%) | 0 (0.0%) |  |
| History of any ischemic<br>event number (%) | 74 (28.8%) | 67<br>(25.9%) | 68<br>(23.0%) | 39<br>(31.7%) | 19<br>(33.9%) | 15<br>(36.6%) | 0.07 |
| **History of ischemic stroke number (%)** | 43 (16.7%) | 47 (18.1%) | 43 (14.5%) | 21 (17.1%) | 14 (25.0%) | 12 (29.3%) | 0.05 |
| **History of peripheral artery disease number (%)** | 19 (7.4%) | 11 (4.2%) | 11 (3.7%) | 13 (10.6%) | 3 (5.4%) | 3 (7.3%) | 0.05 |
| **History of myocardial infarction number (%)** | 26 (10.1%) | 24 (9.3%) | 29 (9.8%) | 12 (9.8%) | 5 (8.9%) | 4 (9.8%) | 1.00 |
| **History of diabetes mellitus number (%)** | 48 (18.7%) | 38 (14.7%) | 38 (12.8%) | 30 (24.4%) | 11 (19.6%) | 7 (17.1%) | 0.03 |
| **History of hypertension number (%)** | 135 (52.7%) | 114 (44.0%) | 123 (41.6%) | 71 (57.7%) | 33 (60.0%) | 22 (53.7%) | <0.01 |
| **INR median (IQR)** | 1.0 (1.0-1.1) | 1.0 (1.0-1.1) | 1.0 (1.0-1.1) | 1.0 (1.0-1.1) | 1.0 (1.0-1.1) | 1.0 (1.0-1.1) | 0.09 |
| **ASPECTS at baseline median (IQR)** | 9 (8-10) | 9 (8-10) | 9 (8-10) | 9 (8-10) | 9 (7-10) | 9 (8-10) | 0.43 |
| **Collateral score number (%)** |  |  |  |  |  |  | 0.39 |
| **0** | 12 (4.7%) | 20 (7.9%) | 22 (7.6%) | 5 (4.1%) | 1 (1.8%) | 4 (9.8%) |  |
| **1** | 79 (31.1%) | 72 (28.3%) | 83 (28.6%) | 36 (29.8%) | 16 (28.6%) | 16 (39.0%) |  |
| **2** | 110 (43.3%) | 105 (41.3%) | 117 (40.3%) | 58 (47.9%) | 27 (48.2%) | 13 (31.7%) |  |
| **3** | 53 (20.9%) | 57 (22.4%) | 68 (23.4%) | 22 (18.2%) | 12 (21.4%) | 8 (19.5%) |  |
| **Occlusion location number (%)** |  |  |  |  |  |  | <0.01 |
| **ICA** | 0 (0.0%) | 3 (1.2%) | 2 (0.7%) | 1 (0.8%) | 0 (0.0%) | 0 (0.0%) |  |
| **ICA-T** | 50 (19.5%) | 63 (24.4%) | 69 (23.4%) | 23 (18.7%) | 11 (19.6%) | 10 (24.4%) |  |
| **M1** | 167<br>(65.0%) | 148<br>(57.4%) | 183<br>(62.0%) | 73<br>(59.3%) | 38<br>(67.9%) | 21<br>(51.2%) |  |
| **M2** | 38 (14.8%) | 41<br>(15.9%) | 41<br>(13.9%) | 26<br>(21.1%) | 5 (8.9%) | 7<br>(17.1%) |  |
| **Other** | 2 (0.8%) | 3 (1.2%) | 0 (0.0%) | 0 (0.0%) | 2 (3.6%) | 3 (7.3%) |  |
| **Time from onset of neurological deficit to randomization in minutes median (IQR)** | 90 (71-151) | 91 (68-134) | 87 (69-134) | 92 (66-149) | 90 (72-128) | 129 (84-171) | 0.05 |
| **Time from onset of neurological deficit to IVT in minutes median (IQR)** | 95 (74-155) | NA | 95 (73-139) | 102 (70-141) | 110 (82-147) | 161 (101-182) | 0.06 |
| **Time from onset of neurological deficit to groin puncture in minutes median (IQR)** | 134<br>(105-184) | 128<br>(103-173) | 127<br>(103-170) | 138<br>(103-180) | 130<br>(109-164) | 161<br>(121-203) | 0.04 |
| **Duration of procedure in minutes median (IQR)** | 45 (30-62) | 49 (33-82) | 46 (30-70) | 50 (36-70) | 43 (32-66) | 52 (33-72) | 0.55 |
| **DL-WML volume in mL median (IQR)** | 2 (0-4) | 2 (0-6) | 1 (0-3) | 3 (1-5) | 6 (2-10) | 24<br>(15-33) | <1e-9 |

### White matter lesion segmentation

**Figure 2**. depicts NCCTs from a patient in the A- and B-test sets. Compared to the A-test set, in the B-test set DSC was lower (B-test median DSC:0.23 IQR:[0.06;0.43] vs. A-test median DSC:0.68 IQR:[0.37;0.77]), and the HD was higher (B-test median HD:32.3 IQR:[26.9;48.3] mm vs. A-test median HD:17.9 IQR:[13.6;23.9] mm). **Figures 3A and 3B** describe the relationship between the number of WML, contrast-to-noise ratio, and DSC. Average volume per WML and the contrast-to-noise ratio were (log)linearly associated with DSC. For a similar average volume per WML and contrast-to-noise ratio both the A- and B-test sets had a similar DSC. Volumetric correspondence between the deep-learning-based segmentations and the ground truth was similar for both test sets in terms of ICC (B-test median ICC:0.91 95%CI:[0.87;0.94] vs. A-test median ICC:0.87 95%CI:[0.71;0.95]) and for the Bland-Altman analysis (B-test bias:-2mL LoA:[-11mL;7mL] vs. A-test bias:-3mL LoA:[-12mL;7mL]) (**Figures 3C and 3D**). For larger volumes, DL-WML underestimated the ground truth volume in both test sets. **Online supplementary Figures S1-6** contain plots relating DSC with all other NCCT quality metrics, results for DL-WML segmentations not using the dilated ventricle mask, and the DL-WML volume per WML-Faz score.

**Figure 2.**
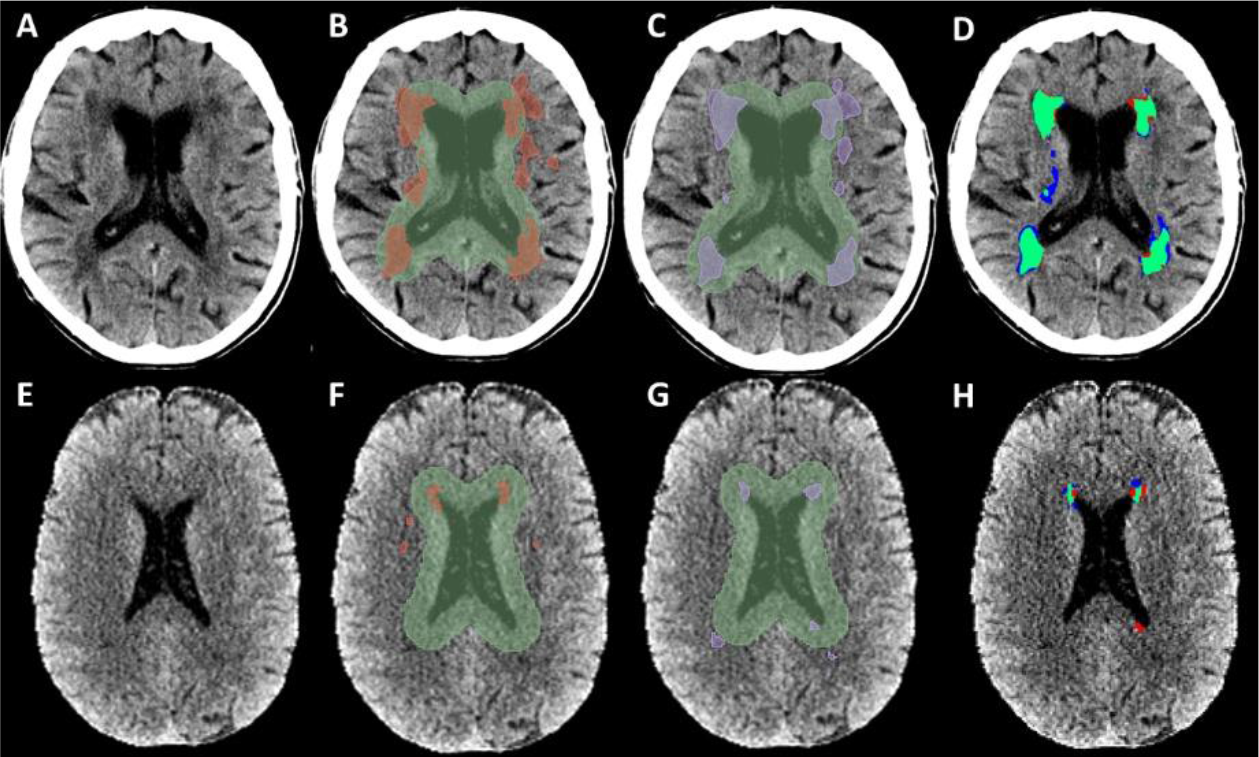
Test set segmentation examples. For anonymization purposes, we received defaced data, with the skull retained for IC data only. For visualization, intensities were clipped between 10 and 60 Hounsfield Units. The dilated ventricle mask (green) was used as a region of interest to exclude false positive segmentations that were not inside the white matter. Only predicted segmentations inside the regions of interest were considered for DL-WML volume measurements. The ground truth values not in the region of interest were not considered for evaluation of the center A- and center B-test set spatial and volumetric correspondence metrics. A/E: The baseline NCCT of a patient from the center A- and center B-test sets. B/F: ground truth segmentation (in red). C/G: DL-WML prediction (in purple). D/H: an overlay with ground truth (red) and predicted (purple) segmentation.

**Figure 3:**
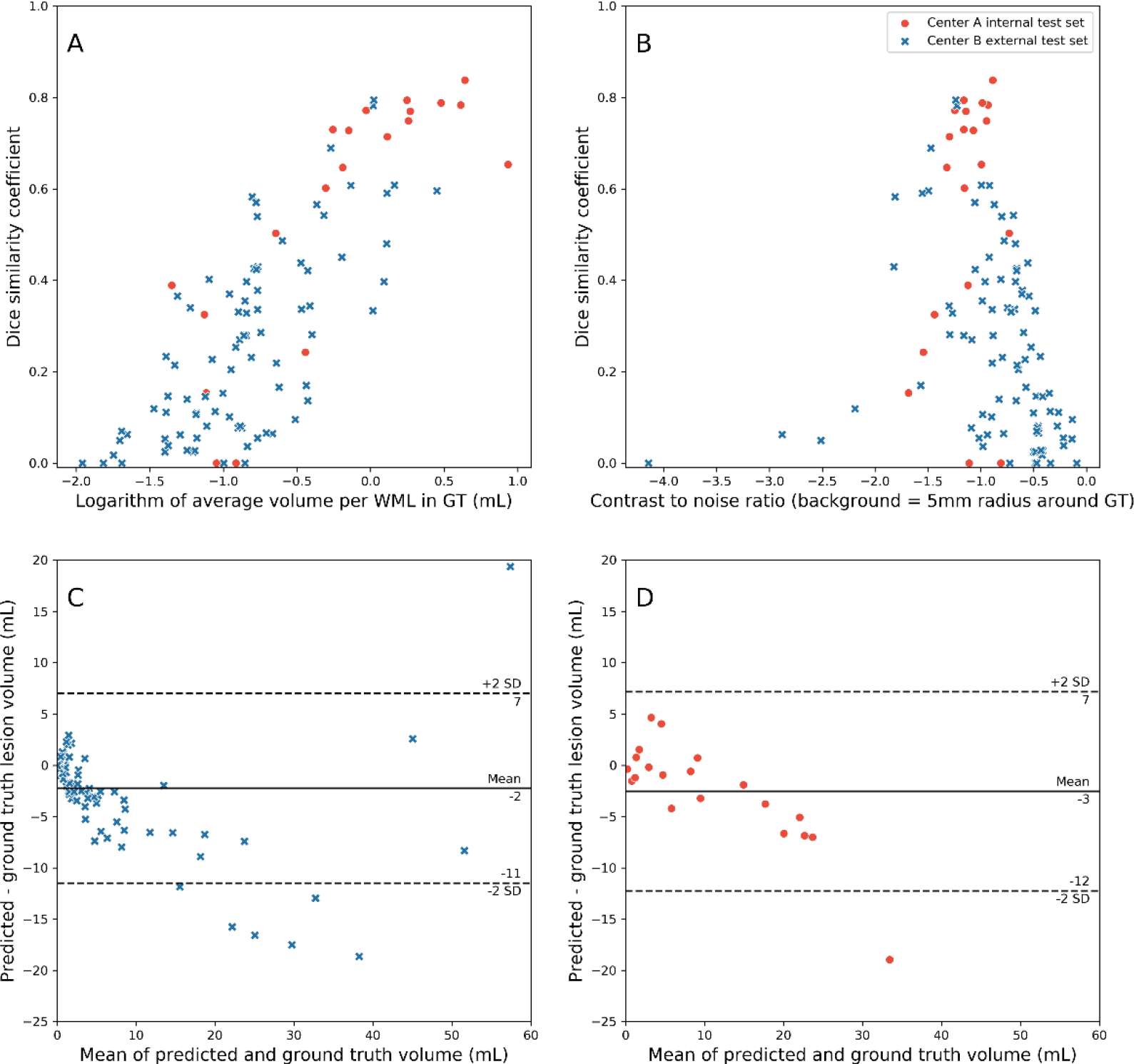
Performance of deep-learning-based WML segmentation compared to ground truth annotations. Results are presented for the center A internal and the center B external test sets. A) Dice similarity coefficient by the logarithm with base 10 of the average WML volume. B) Dice similarity coefficient by contrast-to-noise ratio using a 5mm radius around the ground truth as background. More to the left (more negative) implies more contrast differences between ground truth WML and surrounding brain tissue. C) Bland-Altman plot for the center B-test set. D) Bland-Altman plot for the center A-test set. C-D) Values below zero imply an underestimation of the deep-learning method. GT: ground truth. WML: white matter lesion.

### Post-hoc analysis MR CLEAN No-IV

**Table 3** describes all the associations between WML-Faz and DL-WML volume with the outcome measures without and with adjustment of potential confounders. Both the WML-Faz and DL-WML volume were associated with functional outcome, with and without adjustment for potential confounders for ordinal mRS (DL-WML volume: cOR:0.76[95%CI:0.68;0.83], acOR:0.84[95%CI:0.76;0.94]; WML-Faz: cOR:0.57[95%CI:0.48;0.67], acOR:0.73[95%CI:0.60;0.88]), functional independence (DL-WML volume: cOR:0.65[95%CI:0.55;0.76], acOR:0.76[0.64;0.91]; WML-Faz: cOR:0.51[95%CI:0.41;0.63], acOR:0.62[0.49;0.80]), and sICH (DL-WML volume: cOR:1.26[95%CI:1.08;1.46], acOR:1.31[95%CI:1.08;1.60]; WML-Faz: cOR:1.59[95%CI:1.13;2.24], acOR:1.53[95%CI:1.02;2.31]). WML-Faz and DL-WML volume were both associated with death without adjusting for potential confounders but not after adjusting (DL-WML volume: cOR:1.21[95%CI:1.08;1.46], acOR:1.01[95%CI:0.87;1.17]; WML-Faz: cOR:1.64[95%CI:1.32;2.24], acOR:1.19[95%CI:0.91;1.57]). Neither WML measure was associated with aICH (DL-WML volume: cOR:1.06[95%CI:0.92;1.22], acOR:1.03[95%CI:0.87;1,22]; WML-Faz: cOR:1.24[95%CI:0.99;1.56], acOR:1.16[95%CI:0.91;1.49]). **Online supplementary tables S2-3** contain model fit parameters for all the regression models and a cross-table with sICH for each sub-score on the Fazekas scale. Regression models using DL-WML volume or the WML-Faz had comparable LL for the univariable unadjusted models (ordinal mRS -unadjusted: DL-WML volume LL: 36.14, WML-Faz LR: 45.28; sICH - unadjusted: DL-WML volume LL: 7.23, WML-Faz LR:6.67) and multivariable-adjusted models (ordinal mRS - adjusted: DL-WML volume LL: 189.34, WML-Faz LL: 189.53; sICH - adjusted: DL-WML volume LL: 42.88, WML-Faz LL: 43.98).

**Table 3:**
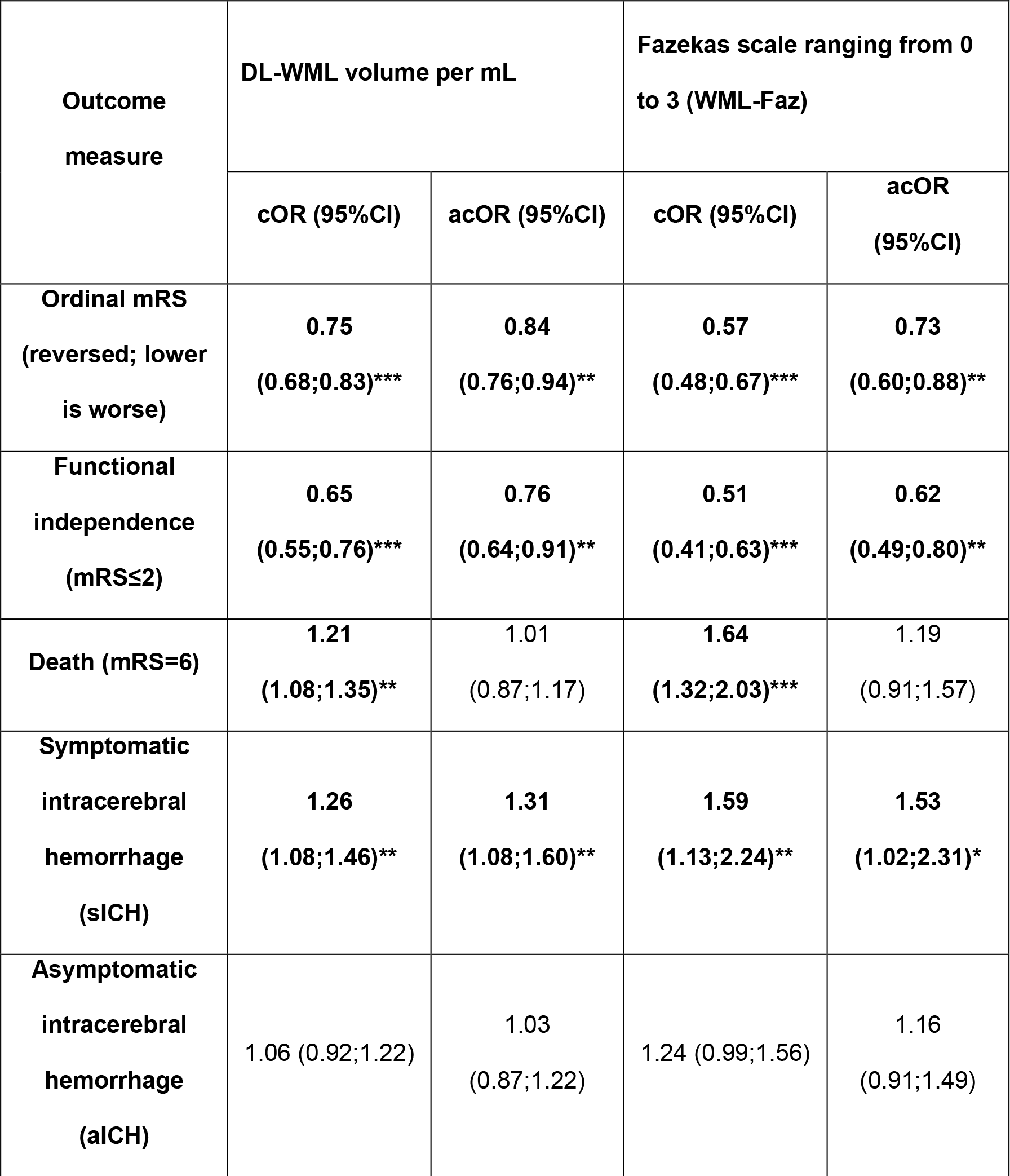
*Association of Fazekas scale and* DL*-WML volume with outcome measures*. Odds ratios are depicted for DL-WML volume per mL and a change in Fazekas scale per point as a result of the regression models. *:<0.05, **:<0.01, ***:<0.0001. DL-WML: deep-learning-based white matter lesion volume. mRS: modified Rankin Scale. cOR: common odds ratio; before adjustment for potential confounders. acOR: adjusted common odds ratio; after adjustment for potential confounders.

| Outcome measure | DL-WML volume per mL |  | Fazekas scale ranging from 0 to 3 (WML-Faz) |  |
| --- | --- | --- | --- | --- |
|  | cOR (95%CI) | acOR (95%CI) | cOR (95%CI) | acOR (95%CI) |
| **Ordinal mRS (reversed; lower is worse)** | **0.75**<br>**(0.68;0.83)\*\*\*** | **0.84**<br>**(0.76;0.94)\*\*** | **0.57**<br>**(0.48;0.67)\*\*\*** | **0.73**<br>**(0.60;0.88)\*\*** |
| **Functional independence (mRS≤2)** | **0.65**<br>**(0.55;0.76)\*\*\*** | **0.76**<br>**(0.64;0.91)\*\*** | **0.51**<br>**(0.41;0.63)\*\*\*** | **0.62**<br>**(0.49;0.80)\*\*** |
| **Death (mRS=6)** | **1.21**<br>**(1.08;1.35)\*\*** | **1.01**<br>**(0.87;1.17)** | **1.64**<br>**(1.32;2.03)\*\*\*** | **1.19**<br>**(0.91;1.57)** |
| **Symptomatic intracerebral hemorrhage (sICH)** | **1.26**<br>**(1.08;1.46)\*\*** | **1.31**<br>**(1.08;1.60)\*\*** | **1.59**<br>**(1.13;2.24)\*\*** | **1.53**<br>**(1.02;2.31)\*** |
| **Asymptomatic intracerebral hemorrhage (aICH)** | 1.06 (0.92;1.22) | 1.03<br>(0.87;1.22) | 1.24 (0.99;1.56) | 1.16<br>(0.91;1.49) |

**Table 4** describes the ORs and p-values for IVT effect modification analyses using an intention-to-treat approach for receiving IVT and EVT. WML-Faz-based IVT effect modification for sICH was statistically significant before adjusting for potential confounders (cOR:8.77 95%CI:[1.04;74.09], p=0.046), but not after adjusting for potential confounders (acOR:7.24 95%CI:[0.71;73.96], p=0.095). DL-WML volume-based IVT effect modification for sICH was not statistically significant (cOR:1.08 95%CI:[0.94;1.25], p=0.274; acOR:1.09 95%CI:[0.93;1.28], p=0.295). Compared to models without WML-Faz-based IVT effect modification for sICH, models with WML-Faz-based IVT effect modification had a higher LL for the unadjusted analyses (LL no WML-Faz-IVT effect modification: 6.67, LL with WML-Faz-IVT effect modification: 11.20) and a similar LL for the adjusted analyses (LL no WML-Faz-IVT effect modification: 43.98, LL with WML-Faz-IVT effect modification: 43.38). For the remainder of outcome measures, no IVT effect modification due to WML-Faz or DL-WML was observed. **Figure 4** depicts the probability of sICH for each Fazekas scale sub-score and by the DL-WML volume for both the intention-to-treat and per-protocol analyses. sICH occurred in 28 (5.4%) and in 25 (5.1%) all patients in the intention-to-treat and per-protocol analysis respectively. **Online supplementary tables S4-6** describe the IVT effect modification results for the per-protocol analysis, a table with model fit statistics, and a cross-table with sICH for each sub-score on the Fazekas scale. The IVT effect modification for sICH due to WML-Faz was higher in the per-protocol analysis and statistically significant with and without adjustment for confounders (cOR: 37.11 95%CI:[3.03;454.13]), p=0.005; acOR:34.52 95%CI:[2.18;545.41], p=0.012). For DL-WML volume-based IVT effect modification for sICH the multiplicative term was non-significant (cOR: 1.20 95%CI:[1.0;1.43]), p=0.052; acOR:1.22 95%CI:[0.99;1.50], p=0.065).

**Table 4:** Associations of multiplicative interaction terms for IVT effect modification with outcome measures. DL-WML: deep-learning-based white matter lesion volume. mRS: modified Rankin Scale. cOR: common odds ratio; before adjustment for potential confounders. acOR: adjusted common odds ratio; after adjustment for potential confounders. *:p<0.05

| Outcome measure | DL-WML volume per mL |  |  |  | Fazekas scale per point between 0 and 3 (WML-Faz) |  |  |  |
| --- | --- | --- | --- | --- | --- | --- | --- | --- |
|  | cOR | p-value | acOR | p-value | cOR | p-value | acOR | p-value |
| **Ordinal mRS (reversed; lower is worse)** | 0.97<br>(0.89;1.05) | 0.441 | 0.98<br>(0.89;1.07) | 0.666 | 1.05<br>(0.4;2.76) | 0.927 | 1.58<br>(0.56;4.49) | 0.389 |
| **Functional independence (mRS≤2)** | 0.99<br>(0.85;1.15) | 0.885 | 1.01<br>(0.87;1.17) | 0.918 | 1.1<br>(0.31;3.91) | 0.881 | 1.59<br>(0.38;6.64) | 0.954 |
| **Death (mRS=6)** | 1.06<br>(0.95;1.17) | 0.308 | 1<br>(0.88;1.13) | 0.993 | 1.8<br>(0.49;6.66) | 0.378 | 0.83<br>(0.17;3.97) | 0.814 |
| **Symptomatic intracerebral hemorrhage (sICH)** | 1.08<br>(0.94;1.25) | 0.274 | 1.09<br>(0.93;1.28) | 0.295 | 8.77<br>(1.04;74.09) | **0.046\*** | 7.24<br>(0.71;73.96) | 0.095 |
| **Asymptomatic intracerebral hemorrhage (aICH)** | 0.98<br>(0.87;1.11) | 0.764 | 0.97<br>(0.86;1.10) | 0.672 | 0.76<br>(0.23;2.55) | 0.662 | 0.59<br>(0.16;2.11) | 0.417 |

**Figure 4:**
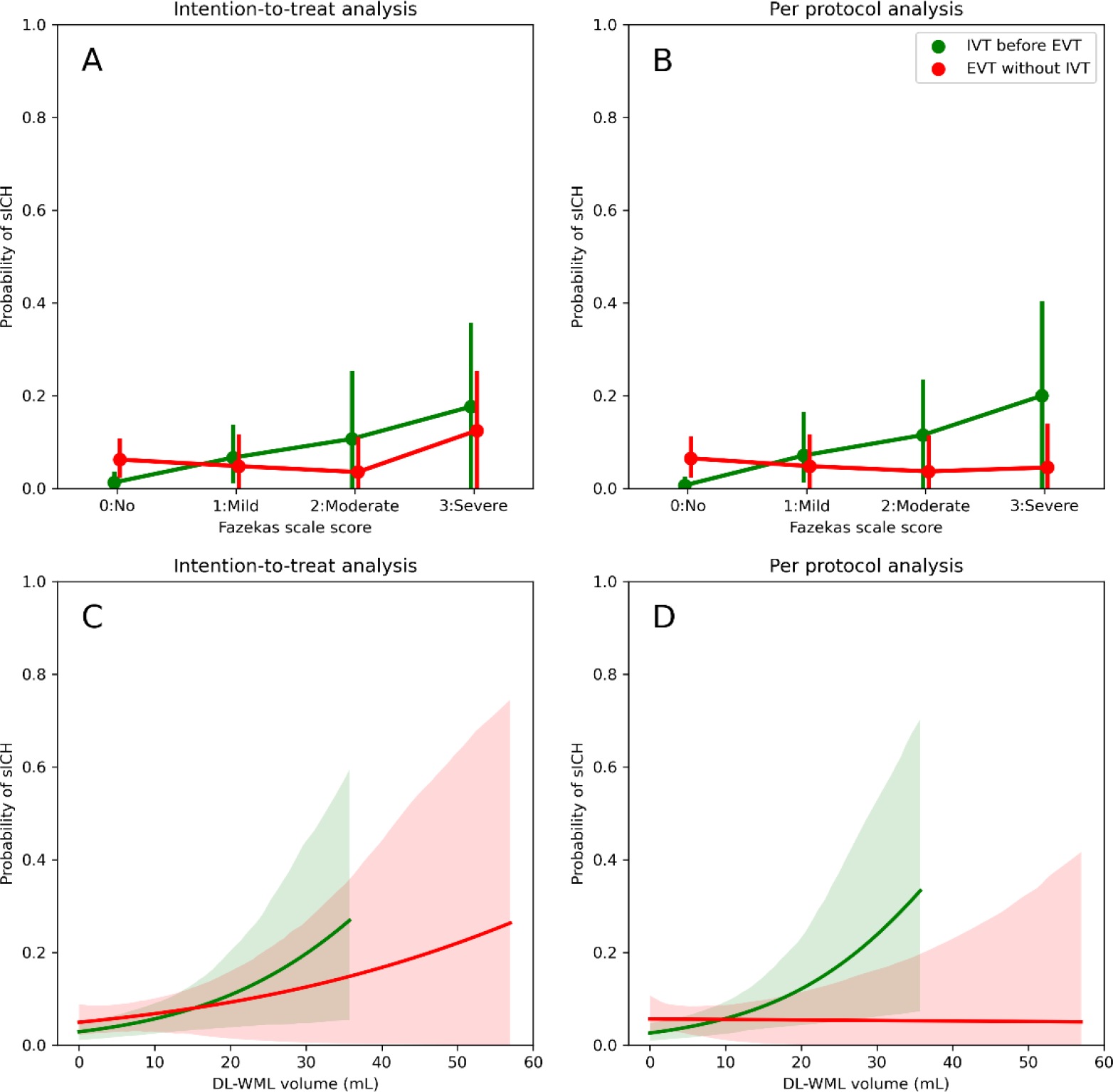
Effect modification for sICH due to IVT and WML load. Mean probability of sICH with 95% confidence interval (error bars / dashed areas) are given. A-B: The x-axis represents sub-scores of WML burden according to the Fazekas scale. A: Probability of sICH by Fazekas scale scores and IVT before EVT considering an intention-to-treat analysis. Unadjusted effect modification p=0.046. B: Probability of sICH by Fazekas scale scores and IVT before EVT considering a per-protocol analysis. Unadjusted effect modification p=0.005. C: Probability of sICH by DL-WML volume and IVT before EVT considering an intention-to-treat analysis. Unadjusted effect modification p=0.274. The green line stops around 35 mL due to absence of data. D: Probabilty of sICH by DL-WML volume for IVT before EVT considering a per-protocol analysis. Unadjusted effect modification p=0.052. For the intention-to-treat analyses, all patients were analyzed as randomized for IVT before EVT. The green line terminates early due to absence of data. In the per-protocol analysis, all patients that did not receive IVT according to the randomization or that did not receive EVT were excluded. sICH: Symptomatic intracerebral hemorrhage. DL-WML volume: deep-learning-based white matter lesion volume. WML-Faz: White matter lesion load according to the Fazekas scale.

## Discussion

An automated deep-learning-based method was developed for the volumetric segmentation of WML in NCCT and validated internally and externally. Differences in DSC between the internal (median DSC:0.68 IQR:[0.37;0.77]) and external (median DSC:0.23 IQR:[0.06;0.43]) validation test sets could be explained by varying contrast-to-noise ratios in the NCCT and smaller WMLs in the external validation test set. The volumetric correspondence between the internal (bias:-3mL LoA:[-12mL;7mL]; mean ICC:) and external (bias:-2mL LoA:[-11mL;7mL]; mean ICC) validation test sets was comparable. DL-WML volumes were lower than the WML volumes (semi-automatically) segmented by experts. This study was the first to show that DL-WML volume and the Fazekas visual rating scale have a similar association with sICH and functional outcome after EVT with or without prior IVT irrespective of patient age and cardiovascular history. We did not find a higher probability of sICH if IVT was given before EVT for patients with a higher WML burden (adjusted interaction: IVT*WML-DL p=0.295 ; IVT*WML-Faz p=0.095).

The post-hoc analyses of the Chinese DIRECT-MT trial only found associations of WML-Faz with functional outcome and not with effect modification for sICH in patients receiving IVT before EVT due to WML-Faz (15). In contrast, our findings suggest that DL-WML volume and WML-Faz were associated with worse functional outcomes and more sICH. This contradiction might be due to differences in cardiovascular risk profiles between the Chinese and Dutch populations (25). Alternatively, the association with mortality was stronger in the DIRECT-MT analyses, potentially indicating missed sICH patients. Furthermore, our regression analyses used WML-Faz as a continuous variable while in the DIRECT-MT analyses the statistical power was reduced by using dichotomized WML-Faz variables (15,26). We observed good volumetric correspondence and statistically significant associations with outcomes even though the spatial correspondence was suboptimal in the external test set compared to the internal test set. Although the performance difference between the internal and external validation test sets could indicate overfitting of our deep-learning model on the training- and internal validation test-sets, this difference was probably related to more patients with few and small WML and lower NCCT quality in the external test set. Specifically, Pitkänen et al. achieved a higher DSC (16), which might be attributed to observed higher average Fazekas score, and thus WML volume, compared to the present work. In line with previous research, the suboptimal DSC could be attributed to difficulties with ground truth segmentation for WML in NCCT (16). Whereas other studies only performed internal validation (16,17), we present an external validation and evaluation of DL-WML volume as a risk factor.

Our study has shortcomings. This study is a post-hoc analysis of a randomized trial with several patients receiving IVT in the group randomized not to receive IVT. In the intention-to-treat analysis, we did not find IVT effect modification due to WML, while in the per-protocol analysis IVT effect modification was present for WML-Faz but not for DL-WML volume. This difference might be due to a subset of patients with a sICH that received IVT after the EVT procedure due to clot-fragmentation in the no IVT arm. Nevertheless, our findings on sICH are based on a small set of patients with sICH. Ideally, a prospective study should be performed selecting patients based on WML for IVT. Lastly, the use of WML volume in the dilated ventricle mask might not fully comprehend the complexity, and thus the etiology, of WML patterns that are more clearly described in some visual WML rating scores (13). Future research should validate the diagnostic accuracy and the associations with sICH and functional outcome of DL-WML volume in a patient-level meta-analysis considering similar studies as the MR CLEAN No-IV trial (12,19–23). Although we reported the contrast-to-noise ratio of an NCCT and the number of WMLs in an NCCT as determinants that affect spatial correspondence, future research should more clearly describe cases where deep-learning-based WML segmentation is reliable or not. Since DL-WML volume is on a continuous scale, it might be used to define a DL-WML volume threshold for excluding patients for IVT before EVT. Such a threshold could prove to be more precise for identifying patients with IVT-induced ICH and poor functional outcome than a threshold based on the Fazekas scale. Other characteristics that can be extracted from WML segmentations such as the number of WML, average WML volume, and WML shapes should be studied for outcome prediction and treatment effect estimation. Finally, automatically extracted WML characteristics could be considered for optimizing anti-platelet and anti-coagulant therapies used as secondary prevention for thrombo-embolic events at the cost of more intracerebral hemorrhages (24,25).

## Data Availability

Data will be made available upon reasonable request to the authors.

https://www.contrast-consortium.nl/

## Abbreviations

AIS: Acute ischemic stroke
NCCT: non-contrast CT
DL-WML volume: deep-learning-based white matter lesion volume
WML-Faz: White matter lesion load according to the Fazekas Scale
IVT: intravenous thrombolysis
EVT: endovascular treatment
sICH/aICH: (a)symptomatic intracerebral hemorrhage
mRS: modified Rankin Scale
DSC: Dice similarity coefficient
HD: Hausdorff distance in mm

## Conclusion

DL-WML volume and WML-Faz had a similar relationship with functional outcome and sICH. Thus, DL-WML volume is a reliable alternative to WML-Faz. A patient-level pooled meta-analysis is required to assess whether DL-WML volume and WML-Faz are associated with a higher probability of sICH and worse functional outcome.

## Acknowledgements

We would like to thank NVIDIA Corporation (Santa Clara, California, USA) for providing a GPU for model development.

## Notes

### Competing Interest Statement

Disclosures and conflicts of interest
Charles Majoie received funds from, CVON/Dutch Heart Foundation and Stryker, (related to this project, paid to institution) and from European Commission, Healthcare Evaluation Netherlands TWIN Foundation (unrelated to this project; all paid to institution) and is shareholder of Nicolab. Bart Emmer received funds from Health Holland Top Sector Life Sciences, Nicolab, Healthcare Evaluation Netherlands (unrelated to this project; all paid to the institution). Henk Marquering is co-founder and shareholder of Nicolab and TrianecT. Alinda Postma received an institutional grant from Siemens healthineers and Bayer Healthcare. Matthan Caan received funds from Health~Holland Top Sector Life Sciences and Nicolab (unrelated to this project; all paid to the institution) and is shareholder of Nicolab. Paul Bentley is funded by the National Institute for Health Research i4i Program II-LA-0814-20007 and the NIHR Imperial Biomedical Research Centre.
The CONTRAST consortium acknowledges the support from the Netherlands Cardiovascular Research Initiative, an initiative of the Dutch Heart Foundation (CVON2015-01: CONTRAST), and from the Brain Foundation Netherlands (HA2015.01.06). The collaboration project is additionally financed by the Ministry of Economic Affairs by means of the PPP Allowance made available by the Top Sector Life Sciences & Health to stimulate public-private partnerships (LSHM17016). This work was funded in part through unrestricted funding by Stryker, Medtronic and Cerenovus. The funding sources were not involved in study design, monitoring, data collection, statistical analyses, interpretation of results, or manuscript writing.

### Funding Statement

Funding:
This research was funded by the Dutch Heart Foundation
(CVON2015-01)

### Author Declarations

The ethics committee of the Amsterdam UMC waived ethical approval for this work reference: W21_509 # 21.561

