## Supplementary material for "Deep-learning-based white matter lesion volume in CT is associated with outcome after acute ischemic stroke": Online Supplement

**Online supplementary material**

### Supplement A: Supplementary methods

*Dataset descriptions*

Data sharing agreements with center A and center B have been signed, local ethical review board approval was granted in accordance with the Helsinki agreement. The MR CLEAN NO-IV was a prospective international multicenter single-blinded randomized controlled trial comparing IVT using 0.9 mg/kg alteplase before EVT with EVT only including 539 patients (1). The study protocol (2) and trial registration (ISRCTN80619088 registered on 31 October 2017) of the MR CLEAN No-IV have been published, institutional review board approval for the trial and its sub-studies have been given, all patients or their legal representatives provided informed consent in accordance with the Helsinki agreement.

*Reuse of data*

Data from center A was included from multiple centers in England. Data from center A existed out of 118 previously published patients with CT that were used to train a machine learning model for Fazekas and WML volume estimation (3).  We added 147 additional patients with CT and used deep-learning-based segmentations to perform spatial and volumetric WML evaluation. Data from center B was included from a single center in Finland. Data from center B has partly been reported before for the spatial and volumetric evaluation of deep-learning-based WML segmentation (4). We retrieved 101/147 patients from center B with CT to perform an external validation of our model. Exclusion of 46 patients occurred due to poor co-registration and the absence of a FLAIR image. We used 516 out of 539 patients from the MR CLEAN No-IV to perform a post-hoc analysis of the risks of IVT before EVT based on WML burden (1). The MR CLEAN No-IV was a randomized controlled trial comparing IVT before EVT with EVT alone and did not study the effect of WML burden (1).

*Center A*

Part of the data has previously been described by Chen et al. (Figure 1 (3)) but was redistributed in our center A- train and test sets (3). 20 patients with NCCT in the center A-test set were randomly sampled with stratification per 10 mL ground truth WML volume from the CT-MRI drawing study in (3). The remaining 40 patients from the CT-MRI drawing study were included together with 58 patients from the CT-drawing study in the center A training set (2 were excluded since they only had a single annotation). Patients in the center A test and center A-1 training set had 2 to 3 expert-based WML annotations that were combined to one ground truth with the Stapler algorithm. All patients included in the center A-2 training set were included retrospectively between 2018 and 2021 from the Imperial College London database for patients with acute ischemic stroke. NCCTs were obtained within 24 hours after symptom onset. Patients in the center A-2 training set had per patient (n=147) annotated slices (number of annotated slices median:11 IQR:7-13). Scans were defaced for anonymization purposes.

*Center B*

Image pairs (NCCT and FLARI) were collected retrospectively from the imaging archive in center B. Part of this data has been previously described (4). The images were screened systemically by a neurologist. Imaging was compromised by various scanners: MRIs were acquired with Siemens and Philips devices and CTs were by Siemens and GE devices. Patients were included if the time interval between MRI and CT sequences was less than 6 months. The images were excluded if they had tumors, cortical infarcts, hematomas, multiple sclerosis lesions, or contusions. The evaluation of the radiologist was noticed. Images were pseudonymized and the skull was removed. All samples were identified by a numerical code and patient-identifiable data was not available to recipient. A strict Material Transfer Agreement was required under the Act on Secondary Use of Health and Social Data. There were 101 image pairs were selected for evaluation.

*Imaging measures*

The Alberta stroke program early CT score (ASPECTS), collateral score, occlusion location, and WML according to the Fazekas scale score at baseline were reported by a dedicated core lab of neuro-radiologists. Every scan was rated once by any of the core-lab members. Although the Fazekas scale was originally developed for MRI, the scores used in this study were based on NCCT.

*Outcome measures*

Functional outcome was measured with the modified Rankin Scale (mRS) 90 days after AIS was assessed through a telephone interview or follow-up outpatient clinic visit. Symptomatic intracranial hemorrhage was defined as neurological deterioration after AIS with an NCCT as an indication that showed an intracranial hemorrhage. Asymptomatic intracranial hemorrhage was found on routine follow-up NCCT 24 hours after AIS that was part of the MR CLEAN No-IV standard imaging protocol.

**Supplement B: Ventricle mask used for selecting the region of interest**

To mask out false positive segmentations that were not in the white matter, we used a 10mm dilated ventricle mask. First, a brain mask was constructed based on thresholding and region growing reported by Sales Barros et al. (3). Inside the brain mask, we made the ventricle mask using a threshold of 17 Hounsfield Units. The top and bottom 40 mm of the brain was removed from the ventricle mask. Subsequently, the connected components larger than 1mm were selected, and holes in the ventricle mask were filled. Finally, a 2D dilation filter with a radius of 10mm was used to construct the region of interest mask. In the analyses the ground truth and predicted WMLs were removed. Thus, only ground truth and predicted WMLs in the 10mm dilated ventricle mask were considered.

| Variable | Included and remainder of No-IV trial | | Number of patients with missing data in the included cohort |
|---|---|---|---|
| | Remainder not included | Included | |
| **Number of patients** | 23 | 516 | |
| **Age median (IQR)** | 71 (54-80) | 71 (62-79) | 0 |
| **Male sex number (%)** | 14 (60.9%) | 291 (56.4%) | 0 |
| **NIHSS at baseline median (IQR)** | 9 (7-12) | 16 (10-20) | 0 |
| **mRS prior to AIS number (%)** | | | 1 |
| **0** | 21 (91.3%) | 353 (68.5%) | |
| **1** | 0 (0.0%) | 100 (19.4%) | |
| **2** | 1 (4.3%) | 48 (9.3%) | |
| **3** | 1 (4.3%) | 10 (1.9%) | |
| **4** | 0 (0.0%) | 4 (0.8%) | |
| **History of any ischemic event number (%)** | 5 (21.7%) | 141 (27.3%) | 0 |
| **History of ischemic stroke number (%)** | 1 (4.3%) | 90 (17.4%) | 0 |
| **History of peripheral artery disease number (%)** | 3 (13.0%) | 30 (5.8%) | 0 |
| **History of myocardial infarction number (%)** | 1 (4.3%) | 50 (9.7%) | 0 |
| **History of diabetes mellitus number (%)** | 4 (17.4%) | 86 (16.7%) | 0 |
| **History of hypertension number (%)** | 11 (47.8%) | 249 (48.3%) | 1 |
| **INR median (IQR)** | 1.0 (1.0-1.1) | 1.0 (1.0-1.1) | 78 |
| **ASPECTS at baseline median (IQR)** | 9 (8-10) | 9 (8-10) | 0 |
| **Collateral score number (%)** | | | 8 |
| **0** | 0 (0.0%) | 32 (6.3%) | |
| **1** | 1 (5.0%) | 151 (29.7%) | |
| **2** | 10 (50.0%) | 215 (42.3%) | |
| **3** | 9 (45.0%) | 110 (21.7%) | |
| **Occlusion location number (%)** | | | 1 |
| **ICA** | 1 (4.3%) | 3 (0.6%) | |
| **ICA-T** | 1 (4.3%) | 113 (21.9%) | |

| | | | |
|---|---|---|---|
| **M1** | 15 (65.2%) | 315 (61.2%) | |
| **M2** | 6 (26.1%) | 79 (15.3%) | |
| **Other** | 0 (0.0%) | 5 (1.0%) | |
| **Time from onset of neurological deficit to randomization in minutes median (IQR)** | | | 0 |
| **Time from onset of neurological deficit to IVT in minutes median (IQR)** | | | 264 |
| **Time from onset of neurological deficit to groin puncture in minutes median (IQR)** | 193 (139-237) | 130 (105-176) | 27 |
| **Duration of procedure in minutes median (IQR)** | 54 (38-85) | 46 (31-70) | 31 |
| **Intention-to-treat intravenous thrombolysis number (%)** | 9 (39.1%) | 257 (49.8%) | 0 |
| **Per-protocol intravenous thrombolysis number (%)** | 9 (39.1%) | 276 (53.5%) | 0 |
| **DL-WML volume in mL median (IQR)** | NA | 2 (0-5) | 0 |

*Online supplementary Table S1. Baseline characteristics of included and not included patients from the MR CLEAN No IV trial.* AIS: acute ischemic stroke. ASPECTS = Alberta Stroke Program Early CT Score. IQR = interquartile range. mRS = modified Rankin Scale. NIHSS = National institute of health stroke. SD = standard deviation. INR: international normalized ratio (secondary hemostasis). DL-WML: deep-learning-based white matter lesion volume. ICA: internal carotid artery. ICA-T: ICA terminus. M1: M1 segment of the middle cerebral artery. M2: M2 segment of the middle cerebral artery. WML: white matter lesion. IVT: intravenous thrombolysis. EVT: endovascular treatment. NA: not available.

| Outcome | Statistical measure | Unadjusted | | Unadjusted interaction | | Adjusted | | Adjusted interaction | |
|---|---|---|---|---|---|---|---|---|---|
| | | DL-WML volume | Fazekas scale | DL-WML volume | Fazekas scale | DL-WML volume | Fazekas scale | DL-WML volume | Fazekas scale |
| Ordinal mRS | Model LL | 36.14 | 45.28 | 37.32 | 45.93 | 189.34 | 189.53 | 189.78 | 190.53 |
| | R2 | 0.07 | 0.09 | 0.07 | 0.09 | 0.32 | 0.32 | 0.32 | 0.32 |
| | adjusted R2 | 0.07 | 0.08 | 0.06 | 0.09 | 0.28 | 0.28 | 0.31 | 0.31 |
| Functional independence (mRS≤2) | Model LL | 40.80 | 45.11 | 41.01 | 45.41 | 158.62 | 158.63 | 161.63 | 162.04 |
| | C-statistic | 0.64 | 0.65 | 0.64 | 0.65 | 0.80 | 0.80 | 0.80 | 0.80 |
| | R2 | 0.10 | 0.11 | 0.10 | 0.11 | 0.35 | 0.35 | 0.36 | 0.36 |
| | adjusted R2 | 0.07 | 0.08 | 0.07 | 0.08 | 0.24 | 0.24 | 0.27 | 0.27 |
| | Brier | 0.230 | 0.229 | 0.230 | 0.229 | 0.183 | 0.183 | 0.182 | 0.181 |
| Death (mRS=6) | Model LL | 10.02 | 19.69 | 14.45 | 23.85 | 129.98 | 129.98 | 131.60 | 131.66 |
| | C-statistic | 0.60 | 0.63 | 0.62 | 0.65 | 0.83 | 0.83 | 0.84 | 0.84 |
| | R2 | 0.03 | 0.06 | 0.05 | 0.07 | 0.36 | 0.36 | 0.37 | 0.37 |
| | adjusted R2 | 0.02 | 0.04 | 0.02 | 0.05 | 0.19 | 0.19 | 0.23 | 0.23 |
| | Brier | 0.146 | 0.143 | 0.145 | 0.142 | 0.105 | 0.105 | 0.105 | 0.105 |
| Symptomatic intracranial hemorrhage (sICH) | Model LL | 7.23 | 6.67 | 8.74 | 11.20 | 42.88 | 43.98 | 40.48 | 43.38 |
| | C-statistic | 0.63 | 0.62 | 0.67 | 0.66 | 0.83 | 0.84 | 0.83 | 0.84 |
| | R2 | 0.04 | 0.04 | 0.05 | 0.06 | 0.23 | 0.24 | 0.22 | 0.23 |
| | adjusted R2 | 0.01 | 0.01 | 0.01 | 0.02 | 0.05 | 0.05 | 0.08 | 0.08 |
| | Brier | 0.050 | 0.050 | 0.050 | 0.050 | 0.045 | 0.045 | 0.046 | 0.045 |
| Asymptomatic intracranial hemorrhage (aICH) | Model LL | 1.24 | 4.68 | 1.64 | 5.05 | 30.33 | 30.73 | 31.47 | 32.14 |
| | C-statistic | 0.55 | 0.57 | 0.54 | 0.57 | 0.64 | 0.64 | 0.65 | 0.65 |
| | R2 | 0.00 | 0.01 | 0.00 | 0.01 | 0.08 | 0.09 | 0.09 | 0.09 |
| | adjusted R2 | 0.00 | 0.01 | 0.00 | 0.01 | 0.08 | 0.06 | 0.09 | 0.06 |
| | Brier | 0.216 | 0.214 | 0.216 | 0.214 | 0.203 | 0.203 | 0.202 | 0.202 |

*Online supplementary Table S2. The goodness of fit statistics per model for the intention-to-treat analyses.* LL: log-likelihood. C-statistic: the area under the receiver operatic characteristic curve. R2: sum of squared errors (approximated for binary outcomes). Adjusted R2: R2 adjusted for use of multiple variables. Brier score: the average difference between the predicted probability and the ground truth class.

| Fazekas scale | sICH + IVT | No sICH + IVT | sICH + no IVT | No sICH + no IVT |
|---|---|---|---|---|
| DL-WML volume <1mL | 2 (16.7%) | 66 (26.7%) | 3 (18.8%) | 70 (28.8%) |
| 0: no WML | 2 (16.7%) | 150 (61.2%) | 9 (56.3%) | 135 (55.8%) |
| 1: Mild WML | 4 (33.3%) | 56 (22.9%) | 3 (18.8%) | 59 (24.4%) |
| 2: Moderate WML | 3 (25%) | 25 (10.2%) | 1 (6.3%) | 27 (11.2%) |
| 3: Severe WML | 3 (25%) | 14 (5.7%) | 3 (18.8%) | 21 (8.7%) |
| Total | 12 | 245 | 16 | 242 |

*Online supplementary Table S3. Intention-to-treat table IVT vs. sICH by Fazekas scale.*
sICH: symptomatic intracerebral hemorrhage. WML: white matter lesion. IVT: intravenous thrombolysis.

| Outcome measure | | DL-WML volume per mL | | | | Fazekas scale per point between 0 and 3 (WML-Faz) | | | |
|---|---|---|---|---|---|---|---|---|---|
| | | cOR | p-value | acOR | p-value | cOR | p-value | acOR | p-value |
| **Ordinal mRS (reversed; lower is worse)** | **OR (95%CI)** | 0.98 (0.9;1.07) | 0.693 | 0.99 (0.9;1.08) | 0.766 | 1.05 (0.38;2.86) | 0.93 | 1.38 (0.47;4.1) | 0.557 |
| **Functional independence (mRS≤2)** | **OR (95%CI)** | 0.99 (0.86;1.15) | 0.936 | 1 (0.86;1.16) | 0.977 | 0.98 (0.27;3.59) | 0.978 | 1.27 (0.3;5.48) | 0.744 |
| **Death (mRS=6)** | **OR (95%CI)** | 1.04 (0.93;1.16) | 0.526 | 0.99 (0.87;1.13) | 0.882 | 1.56 (0.4;6.09) | 0.521 | 0.82 (0.16;4.21) | 0.811 |
| **Symptomatic intracranial hemorrhage (sICH)** | **OR (95%CI)** | 1.2 (1;1.43) | 0.052 | 1.22 (0.99;1.5) | 0.065 | 37.11 (3.03;454.13) | **0.005**** | 34.52 (2.18;545.41) | **0.012*** |
| **Asymptomatic intracranial hemorrhage (aICH)** | **OR (95%CI)** | 0.99 (0.87;1.14) | 0.94 | 1 (0.86;1.15) | 0.963 | 0.91 (0.24;3.52) | 0.892 | 0.74 (0.17;3.25) | 0.691 |

*Online supplementary Table S4. Associations of multiplicative interaction terms for effect modification with outcome measures in the per-protocol analysis.* This table is the per protocol analysis equivalent of table 4 in the main text. DL-WML: deep learning based white matter lesion volume. mRS: modified Rankin Scale. cOR: common odds ratio; before adjustment for potential confounders. acOR: adjusted common odds ratio; after adjustment for potential confounders. *:<0.05, **:<0.01

| Outcome | Statistical measure | Unadjusted | | Unadjusted interaction | | Adjusted | | Adjusted interaction | |
|---|---|---|---|---|---|---|---|---|---|
| | | DL-WML volume | Fazekas scale | DL-WML volume | Fazekas scale | DL-WML volume | Fazekas scale | DL-WML volume | Fazekas scale |
| Ordinal mRS | Model LL | 28.51 | 37.60 | 29.44 | 38.46 | 175.29 | 176.27 | 175.39 | 176.62 |
| | R2 | 0.06 | 0.08 | 0.06 | 0.08 | 0.31 | 0.31 | 0.31 | 0.31 |
| | adjusted R2 | 0.06 | 0.07 | 0.06 | 0.08 | 0.30 | 0.30 | 0.30 | 0.30 |
| Functional independence (mRS≤2) | Model LL | 31.55 | 36.12 | 31.75 | 36.40 | 143.57 | 146.40 | 143.57 | 146.50 |
| | C-statistic | 0.63 | 0.64 | 0.63 | 0.65 | 0.79 | 0.79 | 0.79 | 0.79 |
| | R2 | 0.08 | 0.10 | 0.08 | 0.10 | 0.34 | 0.35 | 0.34 | 0.35 |
| | adjusted R2 | 0.06 | 0.07 | 0.06 | 0.07 | 0.26 | 0.26 | 0.26 | 0.26 |
| | Brier | 0.233 | 0.232 | 0.233 | 0.232 | 0.186 | 0.184 | 0.186 | 0.184 |
| Death (mRS=6) | Model LL | 6.85 | 15.41 | 10.93 | 19.49 | 123.84 | 123.95 | 123.87 | 124.01 |
| | C-statistic | 0.59 | 0.62 | 0.62 | 0.64 | 0.83 | 0.84 | 0.83 | 0.84 |
| | R2 | 0.02 | 0.05 | 0.04 | 0.06 | 0.36 | 0.36 | 0.36 | 0.36 |
| | adjusted R2 | 0.01 | 0.03 | 0.02 | 0.04 | 0.22 | 0.22 | 0.22 | 0.22 |
| | Brier | 0.148 | 0.146 | 0.147 | 0.144 | 0.106 | 0.107 | 0.106 | 0.106 |
| Symptomatic intracranial hemorrhage (sICH) | Model LL | 4.72 | 9.39 | 5.60 | 10.24 | 43.59 | 45.09 | 44.31 | 45.36 |
| | C-statistic | 0.58 | 0.58 | 0.58 | 0.59 | 0.67 | 0.67 | 0.67 | 0.67 |
| | R2 | 0.01 | 0.03 | 0.02 | 0.03 | 0.12 | 0.12 | 0.12 | 0.12 |
| | adjusted R2 | 0.01 | 0.02 | 0.01 | 0.02 | 0.09 | 0.09 | 0.09 | 0.09 |
| | Brier | 0.226 | 0.224 | 0.225 | 0.223 | 0.208 | 0.207 | 0.208 | 0.207 |
| Asymptomatic intracranial hemorrhage (aICH) | Model LL | 3.43 | 4.04 | 7.99 | 13.31 | 36.70 | 36.14 | 40.74 | 43.31 |
| | C-statistic | 0.62 | 0.61 | 0.67 | 0.69 | 0.84 | 0.83 | 0.84 | 0.86 |
| | R2 | 0.02 | 0.02 | 0.05 | 0.08 | 0.22 | 0.21 | 0.24 | 0.26 |
| | adjusted R2 | 0.01 | 0.01 | 0.02 | 0.03 | 0.07 | 0.07 | 0.08 | 0.09 |
| | Brier | 0.048 | 0.048 | 0.048 | 0.047 | 0.044 | 0.044 | 0.042 | 0.042 |

*Online supplementary Table S5.* LL: log-likelihood. C-statistic: the area under the receiver operatic characteristic curve. R2: sum of squared errors (approximated for binary outcomes). Adjusted R2: R2 adjusted for use of multiple variables. Brier score: the average difference between the predicted probability and the ground truth class.

| Fazekas scale | sICH + IVT | No sICH + IVT | sICH + no IVT | No sICH + no IVT |
|---|---|---|---|---|
| DL-WML volume <1mL | 1 (9.1%) | 60 (26.5%) | 3 (21.4%) | 67 (28.4%) |
| 0: no WML | 1 (9.1%) | 139 (61.5%) | 9 (64.3%) | 129 (54.9%) |
| 1: Mild WML | 4 (36.4%) | 52 (23%) | 3 (21.4%) | 59 (25.1%) |
| 2: Moderate WML | 3 (27.3%) | 23 (10.2%) | 1 (7.1%) | 26 (11.1%) |
| 3: Severe WML | 3 (27.3%) | 12 (5.3%) | 1 (7.1%) | 21 (8.9%) |
| Total | 11 | 226 | 14 | 235 |

*Online supplement table S6. Per-protocol analysis table IVT vs. sICH by Fazekas scale.*
sICH: symptomatic intracerebral hemorrhage. WML: white matter lesion. IVT: intravenous thrombolysis.
